# Large Language Models for Structured Information Extraction from German Histopathology Reports – Hepatocellular Carcinoma

**DOI:** 10.64898/2026.10.07.26364402

**Authors:** N. Koch, N. Hartmann, A. Männel, G. Baretton, D. Aust, P. Hönscheid

## Abstract

**Background:** Clinical registries and institutional quality control (QC) in pathology heavily depend on complex histopathological data. However, critical tumor characteristics are mostly locked in unstructured free-text reports. We hypothesize that locally deployed, resource-eWicient language models operating within a secure institutional network can reliably extract relevant variables from German reports of hepatocellular carcinoma (HCC), thereby enabling automated data curation without transferring sensitive patient data.

**Methods:** We developed a local, privacy-preserving open-weight Large Language Model (LLM) pipeline consisting of two consecutive stages: Stage 1 utilizes a lightweight gatekeeper model to isolate the target cohort of confirmed Hepatocellular Carcinoma cases from a screening pool of *N =* 2,864 reports. Stage 2 deploys a high-capacity model to extract 15 predefined histopathological variables, including tumor–node–metastasis staging, histological grading, vascular invasion, and selected immunohistochemical features into structured JSON format. For both Stages, pipeline performance was evaluated using Accuracy, Macro F1-scores, and Cohen’s kappa compared to expert-curated reference annotations.

**Results:** The top-performing Stage 1 gatekeeper isolated the target patient cohort with zero classification errors, effectively reaching complete agreement with the reference standard (ground truth) and correcting inaccurate administrative metadata. For Stage 2 feature extraction, current-generation models demonstrated robust structural reliability, reducing the burden of manual post-processing. The 31B model configuration (Gemma 4) virtually eliminated objective factual errors, achieving near-perfect agreement with the reference standard (Macro F1 > 0.99).

**Conclusion:** Local two-stage pipelines offer a highly accurate, resource-efficient, and secure solution for clinical text processing. This framework serves as a privacy-preserving engine for institutional quality control and the scalable curation of pathology registries.

## Introduction

Hepatocellular carcinoma (HCC) remains a leading cause of cancer-related mortality worldwide, developing across diverse etiologies including viral hepatitis, alcohol consumption, and metabolic liver diseases ^1^. Managing HCC requires standardized diagnostic and therapeutic pathways, where pathology reports contain the decisive tumor characteristics that guide personalized oncological treatment ^2,3^. However, most of this critical information is documented as unstructured free-text within Laboratory Information Systems. This lack of standardization creates a bottleneck for data accessibility ^4^.

Histopathologically, HCC exhibits morphological diversity and is routinely classified using the WHO grading system to determine tumor differentiation ^5,6^. Beyond grading, critical indicators of poor prognosis and post-surgical recurrence include pathological tumor stage (pT), vascular invasion, and surgical resection margin status. Specific immunohistochemical panels are routinely used to confirm hepatocellular differentiation and differentiate early-stage malignant lesions from dysplastic or benign liver lesions ^7,8^. Automated extraction of these complex histopathological parameters is essential for registry curation and retrospective research.

Beyond research applications, structuring routine diagnostic text provides an instrument for institutional quality control and quality assurance. Automated data extraction enables departments to audit records continuously, validate data integrity, and rapidly detect documentation discrepancies.

Traditional natural language processing (NLP) methods often lack the flexibility to accurately capture the semantic nuances and context of complex medical text ^9^. While modern cloud-based Large Language Models (LLMs) demonstrate high extraction performance, transmitting sensitive patient data to external servers raises severe data privacy and ethical concerns. Consequently, there is an urgent clinical need for local AI solutions that can operate entirely within a hospital’s secure network using compact, cost-effective hardware.

### Related Work

Recent studies have demonstrated the feasibility of using LLMs for structuring pathology data. For instance, Truhn et al. (2024) utilized GPT-4 to extract information from English histopathology reports, yet relied on commercial, cloud-based models that raise data privacy concerns ^10^.

Similarly, recent benchmarks evaluated the extraction performance of open-weight models. Wiest et al. (2025) focused on English data, whereas Grothey et al. (2025) processed native German pathology reports ^11,12^. However, these studies typically deployed single-stage extractions on high-end server infrastructure. A scalable, multi-stage pipeline optimized for local deployment on compact consumer hardware has not yet been established.

### Study Objective

In this study, we present a local, privacy-preserving two-stage LLM pipeline for deployment within a secure institutional network. This framework uses an asymmetric design, deploying a lightweight model as a gatekeeper to isolate the target patient cohort from an administrative screening pool, before utilizing a separate, high-capacity model to extract 15 distinct histopathological features. We analyze formatting compliance, required prompt engineering, and error typologies. Finally, we demonstrate the utility of this multi-stage framework as a scalable engine for automated clinical registry curation and institutional quality control.

## Methods

### Study Population and Data Source

This retrospective study utilized an unedited, real-world cohort of all histopathology reports on liver samples containing malignancy generated between 2020 and 2024. Electronic reports were retrieved from the institutional Laboratory Information System (LIS) from the Institute of Pathology at the University Hospital Dresden, Germany. Via SQL queries, we exported an Excel file containing the free text reports and administrative metadata. Text records were subsequently imported into Python for processing. To maintain strict patient data privacy, all text processing was executed with fully pseudonymized datasets.

All extracted pathology reports were composed in native German free-text. The text contains routine German histopathological documentation, characterized by morphological terminology as well as narrative macroscopic and microscopic descriptions.

Each single pathology report in the dataset was finalized by two board-certified pathologists in accordance with the institutional dual-review standard. Over the five-year study period, approximately 15-20 distinct pathologists contributed to the report corpus. This ensures the dataset includes different individual phrasings within a standardized report structure.

An initial dataset of *N* = 2,864 pathology reports involving liver tissue samples was subjected to screening. A subset of 423 cases with histologically confirmed diagnosis of primary Hepatocellular Carcinoma (HCC) was manually identified as the reference standard (Ground Truth). To prevent error propagation from the triage step and isolate the extraction performance of Stage 2, all feature extraction models were evaluated directly against this reference HCC cohort.

## The Two-Stage Framework

To target real-world conditions, and ensure data integrity, we developed a two-stage pipeline that divides the information extraction task into two sequential stages (Figure 1):

**Figure 1:**
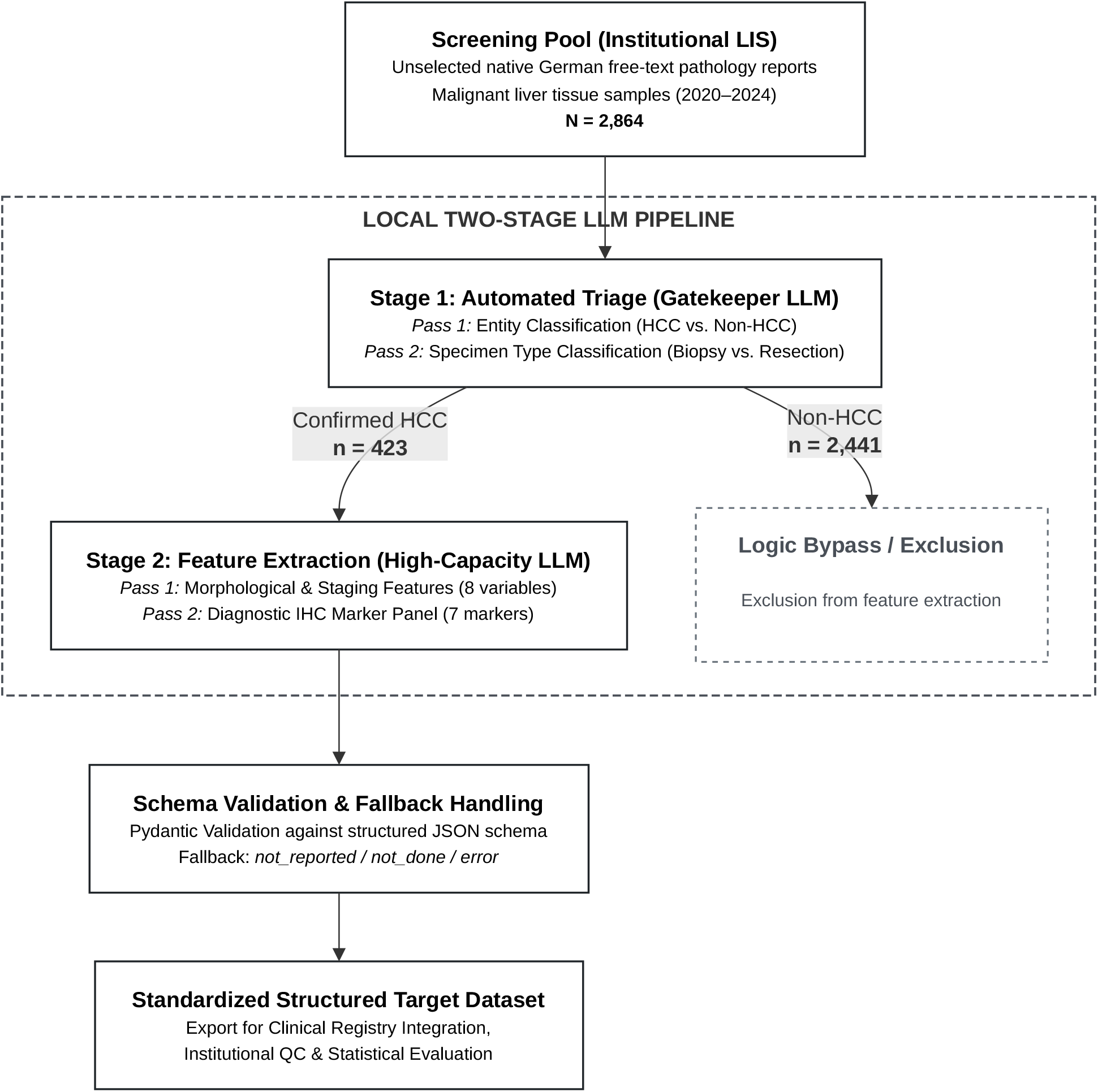
Architecture of the local two-stage Large Language Model (LLM) extraction pipeline, and cohort flow. Schematic overview of the local workflow for unstructured German pathology reports. A screening pool of N = 2,864 unselected routine reports is processed by a lightweight gatekeeper model in Stage 1 to isolate confirmed hepatocellular carcinoma (HCC) cases (n = 423) and correct specimen type while non-HCC cases (n = 2,441) are excluded via a logic bypass. In Stage 2, a high-capacity model extracts 15 predefined features across two consecutive passes (Pass 1: morphological and staging parameters; Pass 2: diagnostic immunohistochemical panel). Extracted outputs undergo Pydantic JSON schema validation and fallback handling.

∘ Stage 1 – Automated Triage (‘Pre-Classification’): This primary filtering layer processes the unselected collection of liver-related reports to exclude non-malignant or unrelated cases and isolate true Hepatocellular Carcinoma (HCC) cases, while simultaneously categorizing the specimen type (biopsy vs. resection).
∘ Stage 2 – Feature Extraction: This downstream stage performs text analysis exclusively on confirmed HCC cases, like those identified in Stage 1. In our study, Stage 2 extracts 15 predefined morphological and immunohistochemical parameters across two sequential passes (Pass 1: 8 morphological and staging parameters; Pass 2: 7 diagnostic IHC markers), required for structured clinical reporting (see Supplementary Tables 1 and 2 for a complete overview of all extracted features, their valid response categories, and reference standard frequencies).

This two-stage approach was specifically introduced to address inconsistencies in legacy metadata: While most entries in the Laboratory Information System (LIS) follow routine indexing, a subset of true HCC cases lacks appropriate top-level administrative tags despite containing the definitive diagnosis within the unstructured text. Relying solely on database queries would therefore result in missed cases. Furthermore, manual auditing revealed that the historical LIS metadata contained a significant error rate regarding specimen type classification (biopsy vs. resection). By evaluating the full-text narrative in Stage 1, the pipeline corrects these database errors and establishes an accurate target cohort before downstream feature extraction.

### Reference Standard / Ground Truth Establishment

To evaluate pipeline performance against a definitive reference standard, a manual annotation process was conducted beforehand to establish the reference cohort. A medical doctor manually screened all 2,864 initial liver reports to validate the Stage 1 triage layer, separating true HCC cases from non-HCC entities as well as deciding biopsy vs resection. For Stage 2 validation, the 423 confirmed HCC reports underwent manual annotation according to the WHO Classification of Tumours (5th Edition, 2019) ^5^. This process captured 8 morphological and staging features (including subtype, pT/pN stage, grading, invasion profiles, and resection status; detailed in Supplementary Table 1) and 7 immunohistochemical (IHC) markers (including Arg-1, CD34, HSP70, GPC3, GS, HepPar-1, and CK7; detailed in Supplementary Table 2).

### Hardware Setup and Backend

All model inferences were executed entirely locally within the secure network infrastructure. The hardware environment consisted of a local workstation equipped with an Apple M2 Pro processor (16-core GPU) and 32 GB of Unified Memory.

Given the strict 32 GB hardware constraint, model compression was mandatory for higher-capacity Stage 2 models to accommodate model weights and KV cache entirely within memory. For these models (27B and 31B), the GGUF framework with advanced block-wise mixed-precision quantization (specifically Q4_K_M) was deployed. Conversely, the compact gatekeeper models evaluated in Stage 1 were operated natively in BF16 precision or using Q8_0 quantization ^13^.

Initially, Ollama was utilized for prototyping due to its abstraction layer, while the native llama.cpp server backend was selected for routine pipeline execution to achieve direct control over hardware allocation and hardware-enforced schema execution ^14,15^.

### Large Language Models

A total of 13 open-weight Large Language Models (LLMs) were screened. While several models were eliminated early due to formatting non-compliance or systematic extraction failures, an iterative prompt-optimization workflow was applied to the models following formatting requirements: For each pass across both Stage 1 and Stage 2, a standard baseline prompt was deployed, evaluated against the reference standard, and manually refined through multi-turn adjustments until peak extraction metrics were reached.

Ultimately, three primary model families were selected for final evaluation and comparative analysis: Ministral 3, Gemma 4, and Qwen 3.5 ^16–18^. To benchmark the performance gains of recent open-weight releases, two previous-generation models – Gemma 3 (27B, Q4_K_M) and GPT-OSS (20B) – were evaluated as baselines across Stage 2 feature extraction. For all evaluations, generation hyperparameters were fixed to greedy decoding (temperature = 0; top_p = 1.0) with a fixed random seed to ensure reproducible outputs.

### Prompt Engineering Design

Prompt configurations utilized a shared, modular blueprint consisting of five distinct blocks: (1) introductory context, (2) a global ruleset, (3) parameter-specific extraction rules, (4) closing constraints, and (5) explicit JSON schema specifications with allowed categories and formatting. All extractions were strictly zero-shot - without providing example reports and corresponding expected answers. Within this universal template, the text depth and inclusion of specific blocks were adjusted dynamically based on the target model.

To ensure linguistic alignment with the source text and avoid transfer errors during inference, all prompts, instructions, and target categories were designed natively in German. Extracted parameters were plotted against English translations of values for analysis and presentation.

### JSON-Output and Schema Validation

To ensure structural consistency, all model inferences utilized the native JSON-mode hyperparameter to strictly enforce JSON-formatted outputs. Once generated, the extracted values and data types were validated against a predefined schema using the Pydantic library to guarantee data integrity.

### Statistical Analysis

Pipeline extraction performance for both stages was quantified using standard classification metrics. To account for feature frequency imbalances within the cohort, the Macro-F1 score was utilized as the primary metric, supplemented by overall accuracy. To quantify the level of agreement between the automated pipeline predictions and the manual reference standard, Cohen’s kappa was calculated for all extracted parameters ^19,20^. To assess the statistical uncertainty and robustness of the model inferences, 95% bootstrap confidence intervals were computed.

All data transformation, evaluation pipelines, and statistical computations were implemented locally using Python.

## Results

### Stage 1: Automated Triage / ‘Pre-Classification’

Stage 1 of the pipeline was evaluated across the entire screening pool (*N* = 2,864) to assess its capability to isolate confirmed Hepatocellular Carcinoma (HCC) cases and classify the underlying specimen types. Overall, the utilized open-weight language models demonstrated high classification performance, as summarized in Table 2 and Supplementary Figures 1 and 2.

**Table 1:** Baseline characteristics of the study cohort. Distribution of the institutional screening pool (N = 2,864) and the confirmed hepatocellular carcinoma (HCC) reference cohort (n = 423) by specimen type (biopsy vs. resection).

| Category - Pathology Reports | Count | Percentage (%) |
| --- | --- | --- |
| Triage Results |  |  |
| - total | 2864 | 100.0% |
| - Confirmed HCC | 423 | 14.8% |
| - Non-HCC | 2441 | 85.2% |
| - of which combined HCC-CCA | 7 | 0.24% |
| Specimen Type (for <u>HCC</u> cohort) |  |  |
| - total | 423 | 100.0% |
| - Biopsy | 248 | 58.6% |
| - Resection | 175 | 41.4% |

**Table 2:** Performance of lightweight gatekeeper models in Stage 1 triage (N = 2,864). Accuracy, macro F1-score and Cohen’s kappa for HCC Status (HCC vs. Non-HCC) and specimen type (biopsy vs. resection) across three open-weight models. Performance is evaluated against the manually annotated institutional reference standard.

| Feature | Model | Accuracy<br>[95% CI] | F1-Score (Macro)<br>[95% CI] | Cohen's Kappa<br>[95% CI] |
| --- | --- | --- | --- | --- |
| HCC Status | Gemma 4 (E4B, BF16) | 1.000 [1.00–1.00] | 1.000 [1.00–1.00] | 1.000 [1.00–1.00] |
|  | Ministral 3 (14B, Q8_0) | 0.999 [1.00–1.00] | 0.998 [1.00–1.00] | 0.996 [0.99–1.00] |
|  | Qwen 3.5 (9B, BF16) | 0.996 [0.99–1.00] | 0.992 [0.99–1.00] | 0.983 [0.97–0.99] |
| Specimen Type<br>(HCC only) | Gemma 4 (E4B, BF16) | 0.993 [0.98–1.00] | 0.993 [0.98–1.00] | 0.985 [0.97–1.00] |
|  | Ministral 3 (14B, Q8_0) | 0.988 [0.98–1.00] | 0.992 [0.98–1.00] | 0.976 [0.95–1.00] |
|  | Qwen 3.5 (9B, BF16) | 0.969 [0.95–0.98] | 0.985 [0.98–0.99] | 0.939 [0.91–0.97] |

*Note: Specimen type classification metrics in Table 2 are computed end-to-end against the entire reference standard HCC cohort ( $n = 423$ ), penalizing triage false negatives as classification errors.*

For the initial triage step (HCC Status), Gemma 4 (E4B, BF16) achieved complete agreement, with accuracy, macro F1-score, and Cohen’s kappa of 1.000 (95% CI: 1.00–1.00). Consequently, the model isolated all 423 true-positive HCC cases without classification errors, whereas the alternative models had misclassifications. In a clinical deployment scenario, even a marginal statistical error rate leads to incomplete registry data downstream.

However, regarding specimen type classification within the isolated HCC cohort, performance remained high across all models, though no model achieved complete agreement with the reference standard. Gemma 4 (E4B, BF16) achieved a macro F1-score of 0.993, closely followed by Ministral 3 (14B, Q8_0) at 0.992 and Qwen 3.5 (9B, BF16) at 0.985.

### Stage 2: Feature Extraction

In Stage 2, the LLM pipeline performed information extraction across 15 distinct histopathological parameters. To benchmark extraction capacities and error boundaries, macro F1-scores along with corresponding 95% bootstrap confidence intervals across all five evaluated models are summarized in Figure 2 and Supplementary Figure 3. Current-generation models markedly outperformed older models (Gemma 3 (27B, Q4_K_M) and GPT-OSS (20B)), which showed substantial degradation in classifications such as grading and histological subtype (Figure 2). The complete metric matrix is provided in Supplementary Table 3.

**Figure 2:**
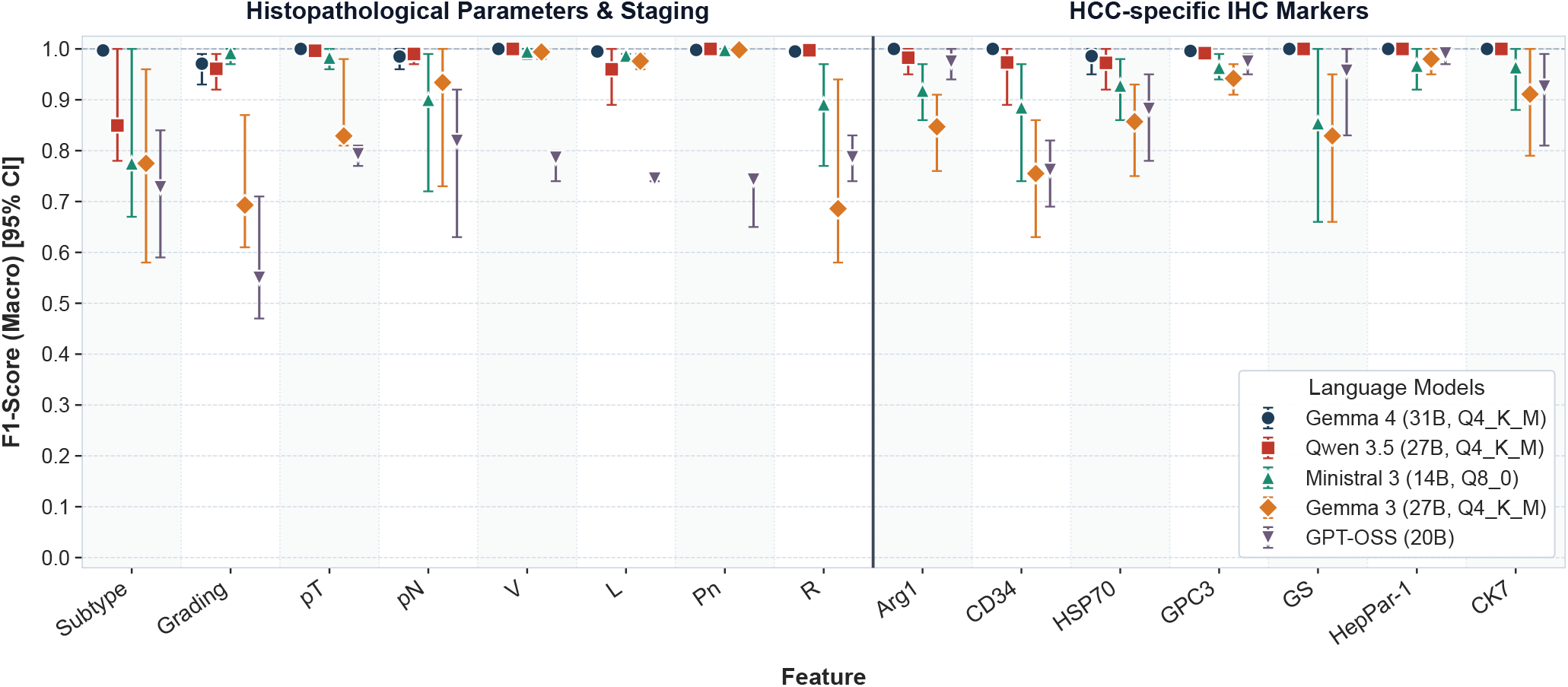
Information extraction performance across 15 histopathological features. Macro F1-scores of open-weight models across the manually curated HCC cohort (n = 423). Features are categorized into morphological characteristics, pathological staging parameters (TNM, grading, resection margins, vascular invasion), and immunohistochemical (IHC) markers. Error bars represent 95% confidence intervals.

Gemma 4 (31B, Q4_K_M) achieved an overall average macro F1-score of 0.993 for morphological tumor features and 0.997 for immunohistochemical (IHC) markers. Notably, Gemma 4 reached an F1-score of 1.000 in critical staging categories including pT stage and vascular invasion (V), as well as in five out of seven IHC markers (Arg-1, CD34, GS, HepPar-1, and CK7).

Qwen 3.5 (27B, Q4_K_M) demonstrated high extraction performance, with an average tumor morphology macro F1-score of 0.969 and an IHC average of 0.989. It matched Gemma 4 in specific sub-parameters, reaching an F1-score of 1.000 for vascular invasion (V), perineural invasion (Pn), as well as for the IHC markers GS, HepPar-1, and CK7.

In contrast, the capacity limits of smaller parameter models became evident with Ministral 3 (14B, Q8_0). While it maintained acceptable performance in standardized classifications like tumor grade (F1 = 0.992) and pT stage (F1 = 0.983), it exhibited significant performance drops in highly variable or contextual parameters. This is indicated by lower macro F1-score for histological subtype extraction (0.775, 95% CI: 0.67–1.00) and specific IHC markers such as CD34 (0.885) and GS (0.854), defining the operational limits of smaller models for reliable clinical data synthesis.

Furthermore, an analysis of absent parameters demonstrated that larger models reliably recognized unrecorded variables without hallucinating speculative values, whereas previous-generation models of comparable scale and smaller models introduced false-positive feature attributions (Supplementary Figure 4).

## Discussion

### Principal Findings

In this study, we demonstrated the clinical feasibility and resource efficiency of a local, asymmetric two-stage Large Language Model (LLM) pipeline for transforming unstructured liver histopathology reports into structured data.

Our asymmetric pipeline design highlights the technical utility of matching task complexity with model scale. For the primary triage layer (Stage 1), the lightweight Gemma 4 (E4B, BF16) configuration functioned as a flawless gatekeeper, isolating all true-positive Hepatocellular Carcinoma (HCC) cases from a screening pool (*N =* 2,864) with zero classification errors. This step addresses an operational bottleneck in clinical text processing: rather than requiring manually pre-filtered cohorts, the pipeline can operate directly on unselected routine data.

Additionally, this high-throughput filtering effectively corrected error-prone administrative tags and specimen type mismatches in legacy Laboratory Information System metadata without deploying high-capacity models across the entire unselected screening cohort. For dense feature extraction (Stage 2), scaling the pipeline up to larger open-weight models (Gemma 4 (31B, Q4_K_M) and Qwen 3.5 (27B, Q4_K_M)) successfully processed the semantic complexity of clinical text.

While preliminary configurations struggled with formatting constraints, all candidate models selected for final benchmarking met the structural requirements, avoiding the formatting failures observed in earlier model generations.

This high reliability reflects recent advances in open-weight Large Language Models: all top-performing models evaluated in this study were the most recent ones, released within six months (Ministral 3 in December 2025, Qwen 3.5 in March 2026, and Gemma 4 in April 2026). Our results show that 4-bit quantized models running on a single 32 GB workstation achieve accurate extraction from German free-text reports without requiring large server clusters.

### Error Typology: Objective Failures vs. Semantic Ambiguity

A detailed review of the discrepancies between model predictions and the manual reference standard revealed that a substantial proportion of recorded variances were not absolute extraction failures, but rather “pseudo-errors” rooted in semantic ambiguity and subjective clinical interpretation. Specifically, out of 18 total deviations recorded for Gemma 4 across all 15 parameters, 16 were subjective: in these instances, both the LLM’s structured output and the human annotator’s reference classification could be considered clinically defensible based on the unstructured narrative text of the original pathology report.

Qwen 3.5 (27B, Q4_K_M) and Gemma 4 (31B, Q4_K_M) committed an identical number of these interpretation-based pseudo-errors, demonstrating comparable ability to interpret complex medical syntax. However, the critical performance divergence lies in the occurrence of true, objective extraction errors. While Qwen 3.5 still introduced occasional objective mis-extractions (such as factual parameter mismatches), Gemma 4 eliminated nearly all true extraction errors, with only two objective classification discrepancies remaining. This finding indicates that Gemma 4 is the more reliable model for autonomous clinical deployment, as it preserves high-level interpretive flexibility while mitigating factual failures in our workflow.

### Model-Specific Prompt Dependency

While the core prompt configuration remained uniform, optimal execution required model-specific engineering. Extraction performance depended primarily on the level of detail and text length in each prompt block. For example, the highly detailed constraints that optimized performance in Qwen 3.5 caused context errors when applied to Gemma 4, and vice versa. Therefore, while a universal template serves as a starting point, clinical deployment requires model-specific tuning to match the processing characteristics of each model family.

### Future Directions

While this study validates the workflow for HCC, scaling the framework to other malignancies presents new opportunities and biological challenges. Immediate expansions should target intrahepatic cholangiocarcinoma (iCCA) and liver metastases.

For this purpose, the pipeline will pivot to an iCCA vs. non-iCCA classification paradigm, with ‘non-iCCA’ serving as a collective placeholder for the remaining heterogeneous entities. For this new task, Stage 1 triage is predicted to face increased classification diWiculty due to different levels of diagnostic certainty (such as possible, probable, or confirmed) instead of simple binary classifications. As iCCA presents histologically as an adenocarcinoma, its narrative descriptions heavily overlap with secondary metastatic adenocarcinomas from extrahepatic primaries.

Differentiating these entities within unstructured text requires the Stage 1 gatekeeper to acknowledge probability, suggesting that a simple binary decision-making approach may no longer suffice.

Beyond hepatic pathology, this asymmetric two-stage framework – utilizing a lightweight gatekeeper for cohort isolation followed by a larger model for feature extraction – is highly generalizable to other complex oncological domains (e.g., breast, lung, or prostate cancer), using tissue-specific prompt matrices ^21^. Furthermore, this workflow is not limited to oncology; it can be scaled to extract data from any complex, unstructured narrative text across pathology and general clinical medicine.

## Data Availability

All data produced in the present study are available upon reasonable request to the authors.

## Competing Interest Statement

The authors declare that they have no competing financial or personal interests that could inappropriately influence or bias the work presented in this manuscript.

## Funding Statement

none

## Ethical Approval

All patients have consented to the release of samples for later use in diagnostic improvement prior to resection or biopsy. In assessing this ethics vote, approval was obtained from the ethics committee of the Medical Faculty of Dresden University of Technology which bases its decision on the guidelines of the World Medical Association in Helsinki in the currently valid version for ‘ICH-GCP-E6’.

## Data Availability Statement

The clinical and histopathological datasets analyzed during the current study contain sensitive patient information protected under GDPR regulations and are not publicly available. De-identified data extracts may be made available from the corresponding author upon reasonable academic request and subject to institutional data transfer agreements.

## Notes

### Competing Interest Statement

The authors have declared no competing interest.

### Author Declarations

All patients gave their informed consent prior to resection or biopsy. In assessing this ethics vote, approval was obtained from the ethics committee of the Medical Faculty of Dresden University of Technology based on the currently valid version for 'ICH-GCP-E6'.

