## Supplementary Material for "Large Language Models for Structured Information Extraction from German Histopathology Reports – Hepatocellular Carcinoma"

*Supplementary Table 1: Reference standard distribution of morphological and staging parameters in the HCC cohort (n = 423).*

| Category | Feature / Parameter | count | Percentage (%) |
|---|---|---|---|
| | - total | 423 | 100.00% |
| Histologic Subtype | NOS | 373 | 88.18% |
| | Clear cell | 16 | 3.78% |
| | Lymphocyte-rich | 1 | 0.24% |
| | Macrotrabecular-massive | 2 | 0.47% |
| | Neutrophil-rich | 1 | 0.24% |
| | Steatohepatitic | 29 | 6.86% |
| | Scirrhous | 1 | 0.24% |
| pT-Stage | pT1a | 25 | 5.91% |
| | pT1b | 68 | 16.08% |
| | pT2 | 43 | 10.17% |
| | pT3 | 20 | 4.73% |
| | pT4 | 12 | 2.84% |
| | not_reported | 255 | 60.28% |
| pN-Stage | pN0 | 27 | 6.38% |
| | pN1 | 2 | 0.47% |
| | pNx | 140 | 33.10% |
| | not_reported | 254 | 60.05% |
| Grading (WHO) | G1 | 147 | 34.75% |
| | G2 | 221 | 52.25% |
| | G3 | 36 | 8.51% |
| | G4 | 1 | 0.24% |
| | Gx | 4 | 0.95% |
| | not_reported | 14 | 3.31% |
| Vascular Invasion | V0 | 160 | 37.83% |
| | V1 | 27 | 6.38% |
| | V2 | 3 | 0.71% |
| | not_reported | 233 | 55.08% |
| Lymphatic Invasion | L0 | 162 | 38.30% |
| | L1 | 10 | 2.36% |
| | not_reported | 251 | 59.34% |
| Perineural Invasion | Pn0 | 179 | 42.32% |
| | Pn1 | 2 | 0.47% |
| | not_reported | 242 | 57.21% |
| Resection Status | R0 | 161 | 38.06% |
| | R1 | 7 | 1.65% |
| | Rx | 6 | 1.42% |
| | not_reported | 249 | 58.87% |

*Supplementary Table 2: Reference standard distribution of the diagnostic immunohistochemical (IHC) marker panel in the HCC cohort (n = 423).*

| IHC | Status | count | Percentage (%) |
|---|---|---|---|
| | - total | 423 | 100.00% |
| Arginase 1 | positive | 154 | 36.41% |
| | negative | 22 | 5.20% |
| | not_done | 247 | 58.39% |
| CD34 | positive | 264 | 62.41% |
| | negative | 8 | 1.89% |
| | not_done | 151 | 35.70% |
| HSP70 | positive | 157 | 37.12% |
| | negative | 12 | 2.84% |
| | not_done | 254 | 60.05% |
| GPC3 | positive | 134 | 31.68% |
| | negative | 57 | 13.48% |
| | not_done | 232 | 54.85% |
| GS | positive | 174 | 41.13% |
| | negative | 4 | 0.95% |
| | not_done | 254 | 60.05% |
| HEPPAR | positive | 58 | 13.71% |
| | negative | 21 | 4.96% |
| | not_done | 344 | 81.32% |
| CK7 | positive | 7 | 1.65% |
| | negative | 94 | 22.22% |
| | not_done | 322 | 76.12% |

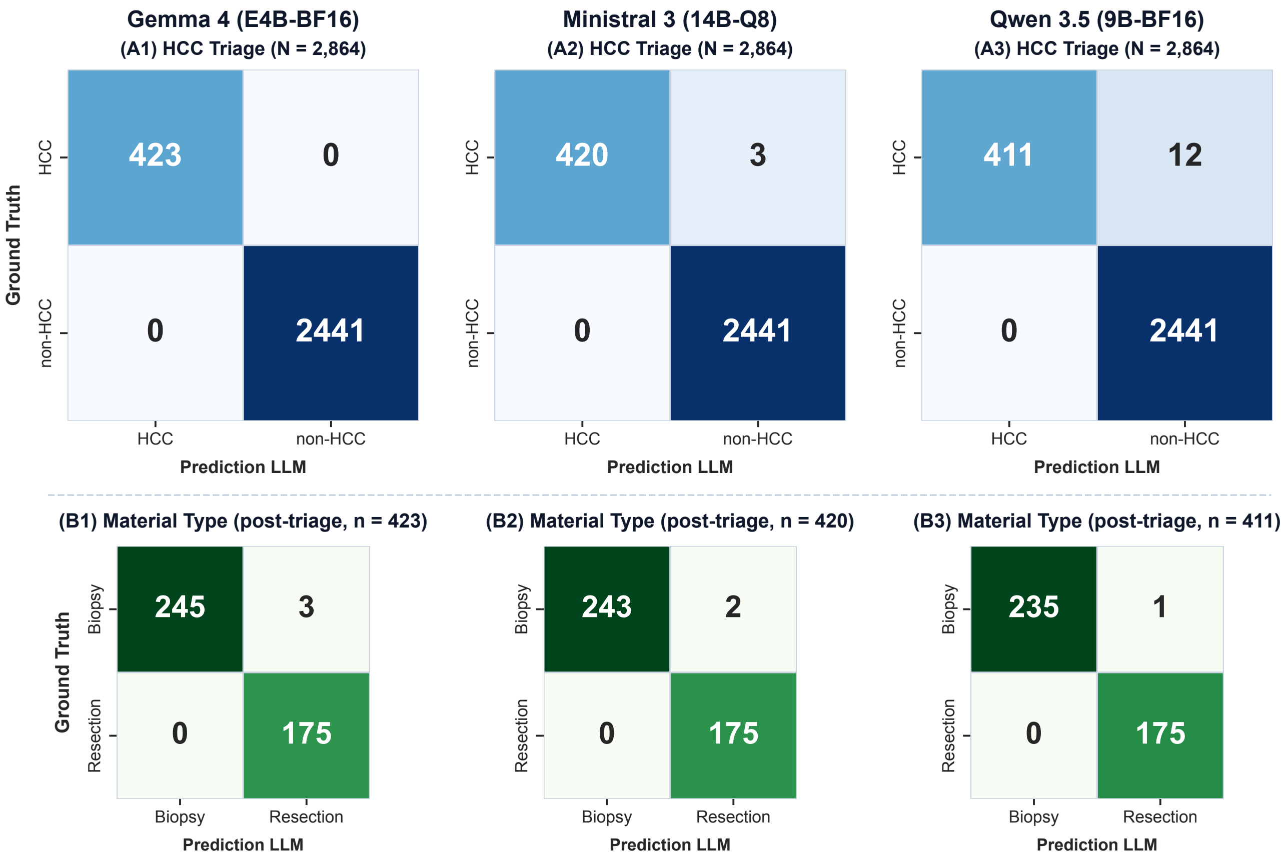


*Supplementary Figure 1: Confusion matrices for Stage 1 triage (HCC entity) and specimen type classification (biopsy vs. resection) across evaluated models.*

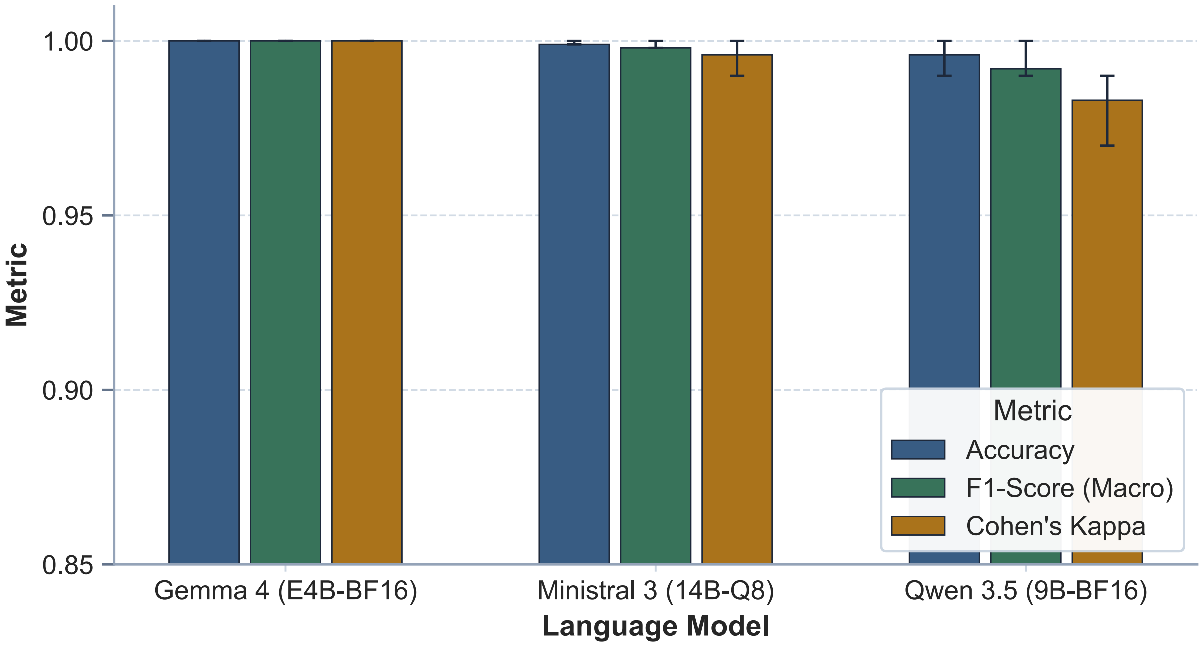


*Supplementary Figure 2: Stage 1 triage performance across evaluated gatekeeper models.*

*Supplementary Table 3: Extraction performance of top-performing Stage 2 models across 15 histopathological features (n = 423). Accuracy, macro F1-score, and Cohen's kappa, 95% confidence intervals for Gemma 4 (31B, Q4_K_M), Qwen 3.5 (27B, Q4_K_M), and Ministral 3 (14B, Q8_0).*

| | Gemma 4 (31B, Q4_K_M) | | | Qwen 3.5 (27B, Q4_K_M) | | | Ministral 3 (14B, Q8_0) | | |
|---|---|---|---|---|---|---|---|---|---|
| | Accuracy [95% CI] | F1-Score (Macro) [95% CI] | Cohen's Kappa [95% CI] | Accuracy [95% CI] | F1-Score (Macro) [95% CI] | Cohen's Kappa [95% CI] | Accuracy [95% CI] | F1-Score (Macro) [95% CI] | Cohen's Kappa [95% CI] |
| Feature | | | | | | | | | |
| Subtype | 0.998 [0.99–1.00] | 0.997 [0.99–1.00] | 0.989 [0.96–1.00] | 0.993 [0.98–1.00] | 0.850 [0.78–1.00] | 0.967 [0.93–1.00] | 0.993 [0.98–1.00] | 0.775 [0.67–1.00] | 0.968 [0.92–1.00] |
| Grading | 0.991 [0.98–1.00] | 0.971 [0.93–0.99] | 0.984 [0.97–1.00] | 0.984 [0.97–0.99] | 0.961 [0.92–0.99] | 0.972 [0.95–0.99] | 0.993 [0.98–1.00] | 0.992 [0.97–1.00] | 0.988 [0.97–1.00] |
| pT | 1.000 [1.00–1.00] | 1.000 [1.00–1.00] | 1.000 [1.00–1.00] | 0.998 [0.99–1.00] | 0.996 [0.99–1.00] | 0.996 [0.99–1.00] | 0.988 [0.98–1.00] | 0.983 [0.96–1.00] | 0.980 [0.96–1.00] |
| pN | 0.993 [0.98–1.00] | 0.985 [0.96–1.00] | 0.987 [0.97–1.00] | 0.995 [0.99–1.00] | 0.990 [0.97–1.00] | 0.991 [0.98–1.00] | 0.988 [0.98–1.00] | 0.900 [0.72–0.99] | 0.978 [0.96–1.00] |
| V | 1.000 [1.00–1.00] | 1.000 [1.00–1.00] | 1.000 [1.00–1.00] | 1.000 [1.00–1.00] | 1.000 [1.00–1.00] | 1.000 [1.00–1.00] | 0.998 [0.99–1.00] | 0.995 [0.98–1.00] | 0.996 [0.99–1.00] |
| L | 0.993 [0.98–1.00] | 0.995 [0.99–1.00] | 0.986 [0.97–1.00] | 0.993 [0.98–1.00] | 0.960 [0.89–1.00] | 0.986 [0.97–1.00] | 0.981 [0.97–0.99] | 0.987 [0.98–0.99] | 0.963 [0.94–0.99] |
| Pn | 0.998 [0.99–1.00] | 0.998 [0.99–1.00] | 0.995 [0.99–1.00] | 1.000 [1.00–1.00] | 1.000 [1.00–1.00] | 1.000 [1.00–1.00] | 0.998 [0.99–1.00] | 0.998 [0.99–1.00] | 0.995 [0.99–1.00] |
| R | 0.991 [0.98–1.00] | 0.995 [0.99–1.00] | 0.981 [0.96–1.00] | 0.995 [0.99–1.00] | 0.997 [0.99–1.00] | 0.991 [0.98–1.00] | 0.979 [0.96–0.99] | 0.891 [0.77–0.97] | 0.959 [0.93–0.99] |
| arg1 | 1.000 [1.00–1.00] | 1.000 [1.00–1.00] | 1.000 [1.00–1.00] | 0.995 [0.99–1.00] | 0.983 [0.95–1.00] | 0.991 [0.98–1.00] | 0.981 [0.97–0.99] | 0.918 [0.86–0.97] | 0.963 [0.94–0.99] |
| cd34 | 1.000 [1.00–1.00] | 1.000 [1.00–1.00] | 1.000 [1.00–1.00] | 0.998 [0.99–1.00] | 0.973 [0.89–1.00] | 0.995 [0.98–1.00] | 0.991 [0.98–1.00] | 0.885 [0.74–0.97] | 0.980 [0.96–1.00] |
| hsp70 | 0.998 [0.99–1.00] | 0.986 [0.95–1.00] | 0.995 [0.99–1.00] | 0.995 [0.99–1.00] | 0.973 [0.92–1.00] | 0.991 [0.98–1.00] | 0.988 [0.98–1.00] | 0.928 [0.86–0.98] | 0.976 [0.95–1.00] |
| gpc3 | 0.998 [0.99–1.00] | 0.996 [0.99–1.00] | 0.996 [0.99–1.00] | 0.995 [0.99–1.00] | 0.992 [0.98–1.00] | 0.992 [0.98–1.00] | 0.979 [0.96–0.99] | 0.963 [0.94–0.99] | 0.963 [0.94–0.99] |
| gs | 1.000 [1.00–1.00] | 1.000 [1.00–1.00] | 1.000 [1.00–1.00] | 1.000 [1.00–1.00] | 1.000 [1.00–1.00] | 1.000 [1.00–1.00] | 0.993 [0.98–1.00] | 0.854 [0.66–1.00] | 0.986 [0.97–1.00] |
| heppar | 1.000 [1.00–1.00] | 1.000 [1.00–1.00] | 1.000 [1.00–1.00] | 1.000 [1.00–1.00] | 1.000 [1.00–1.00] | 1.000 [1.00–1.00] | 0.993 [0.98–1.00] | 0.967 [0.92–1.00] | 0.978 [0.95–1.00] |
| ck7 | 1.000 [1.00–1.00] | 1.000 [1.00–1.00] | 1.000 [1.00–1.00] | 1.000 [1.00–1.00] | 1.000 [1.00–1.00] | 1.000 [1.00–1.00] | 0.988 [0.98–1.00] | 0.964 [0.88–1.00] | 0.969 [0.94–0.99] |
| Mean Tumor | 0.995 | 0.993 | 0.990 | 0.995 | 0.969 | 0.988 | 0.990 | 0.940 | 0.978 |
| Mean IHC | 0.999 | 0.997 | 0.999 | 0.998 | 0.989 | 0.996 | 0.988 | 0.926 | 0.974 |

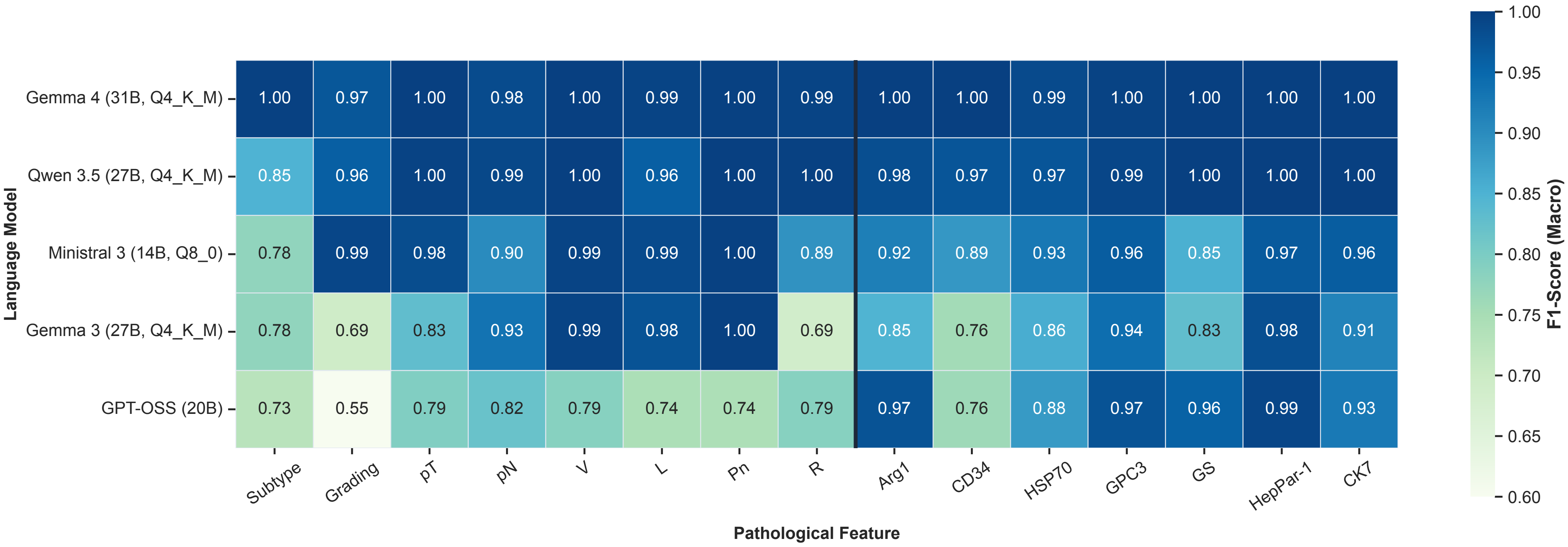


*Supplementary Figure 3: Heatmap of macro F1-scores across all 15 histopathological features for five evaluated local models in Stage 2.*

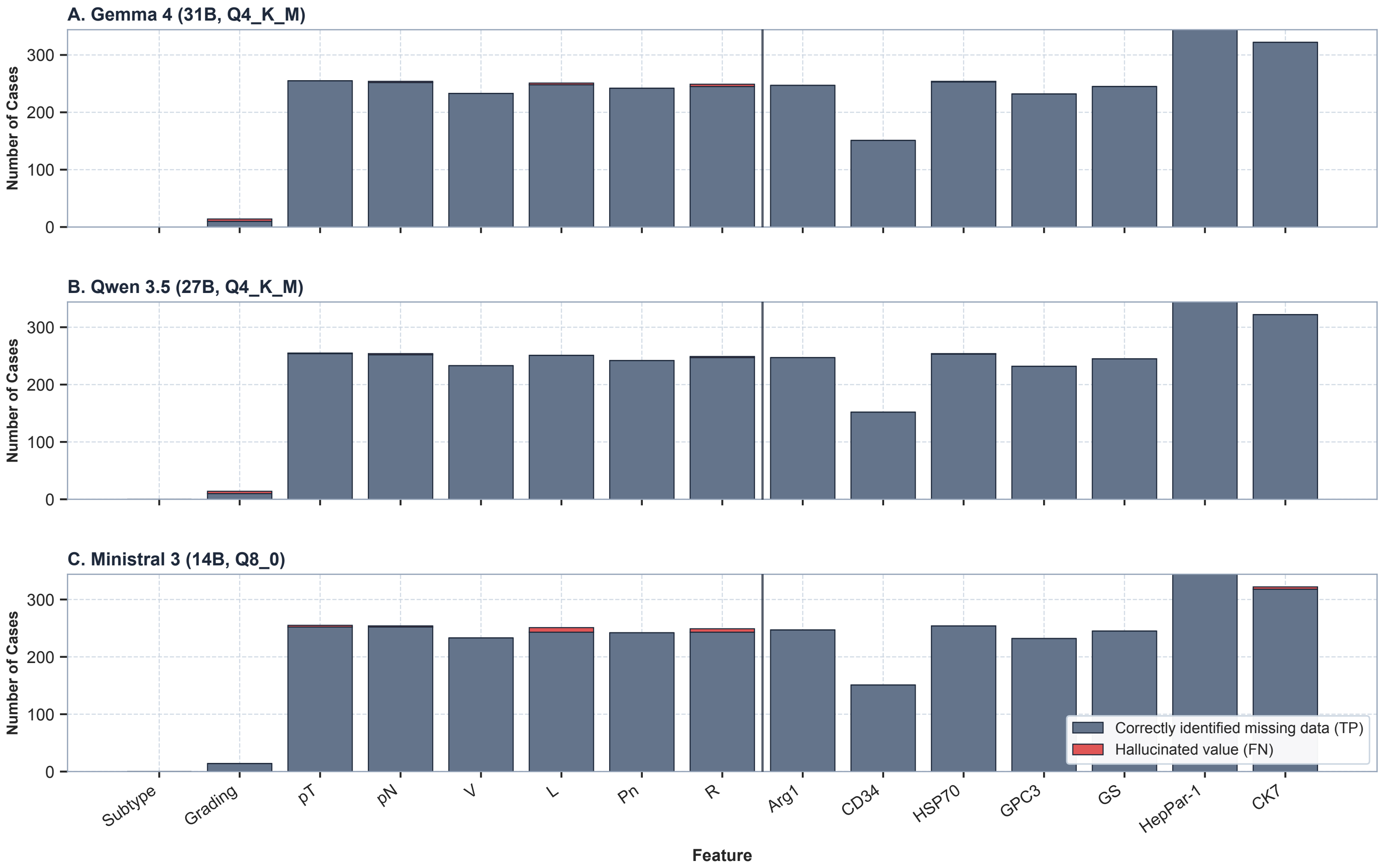


*Supplementary Figure 4: Identification of unrecorded features across top-performing Stage 2 models. Correctly recognized missing histopathological parameters (not_reported / not_done) versus false extractions (hallucinations) across Gemma 4 (31B, Q4_K_M), Qwen 3.5 (27B, Q4_K_M), and Ministral 3 (14B, Q8_0)*
